# microRNA signatures of perioperative myocardial injury after elective non-cardiac surgery: prospective observational cohort study

**DOI:** 10.1101/2020.02.24.20027383

**Authors:** Shaun M. May, Tom E.F. Abbott, Ana Gutierrez Del Arroyo, Anna Reyes, Gladys Martir, Robert C.M. Stephens, David Brealey, Brian H. Cuthbertson, Duminda N. Wijeysundera, Rupert M. Pearse, Gareth L. Ackland

## Abstract

**Background:** Elevated plasma/serum troponin, indicating perioperative myocardial injury (PMI), is common after non-cardiac surgery. However, underlying mechanisms remain unclear. Acute coronary syndrome (ACS) is associated with the early appearance of circulating microRNAs, which regulate post-translational gene expression. We hypothesised that if PMI and ACS share pathophysiological mechanisms, common microRNA signatures should be evident.

**Methods:** Nested case-control study of samples obtained before and after non-cardiac surgery from patients enrolled in two prospective observational studies of PMI (postoperative troponin I/T>99^th^ centile). In cohort one, serum microRNAs were compared between patients with/without PMI, matched for age, gender and comorbidity. Real-time polymerase chain reaction quantified relative microRNA expression (cycle quantification threshold <37) before and after surgery for microRNA signatures associated with ACS, blinded to PMI. In cohort two, we analysed (EdgeR) microRNA from plasma extracellular vesicles using next-generation sequencing (Illumina HiSeq500). microRNA-messenger RNA-function pathway analysis was performed (DIANA miRPath v3.0/TopGO).

**Results:** MicroRNA were detectable in all 59 patients (median age:67yrs (61-75); 42% male), who had similar clinical characteristics independent of developing PMI. In cohort one, PMI was not associated with increased serum microRNA expression levels after surgery (hsa-miR-1-3p mean fold-change (FC):3.99 (95%CI:1.95-8.19); hsa-miR-133-3p FC:5.67(95%CI:2.94-10.91); p<0.001). hsa-miR-208b-3p was more commonly detected after PMI (odd ratio (OR):10.0 (95%CI:1.9-52.6); p<0.01). Bioinformatic analysis of differentially expressed microRNAs from cohorts one and two identified pathways associated with adrenergic stress involving calcium dysregulation, rather than ischaemia.

**Conclusions:** Circulating microRNAs synonymous with cardiac ischaemia were universally elevated in patients after surgery, independent of developing myocardial injury.

## Introduction

Perioperative myocardial injury, as defined by elevated plasma troponin after non cardiac surgery occurs in up to ≥25% patients^1^ and precedes subsequent non-cardiac morbidity.^2^ Preoperative computerized tomography coronary angiogram shows that perioperative myocardial injury occurs across the spectrum of coronary artery disease, suggesting that there are likely to be several pathophysiologic mechanisms that contribute to asymptomatic elevations in troponin after surgery.^3^

Acute coronary syndrome (ACS) is associated with the release of microRNAs from cardiac tissue, frequently before increases in plasma troponin are detectable.^4^ MicroRNAs are small non-coding RNA molecules (approximately 23 nucleotides) that modulate gene expression through the degradation of messenger RNA and/or inhibit messenger RNA translation.^5^ MicroRNAs regulate cardiomyocyte differentiation and survival, calcium regulation, apoptosis, conduction, inflammation and necrosis.^6^ MicroRNAs also regulate intercellular communication via extracellular vesicles released from surface of the cell membrane during activation and/or exosomes, which are smaller luminal vesicles (30– 100 nm in diameter) originating from intracellular endosomes.^7^ MicroRNA-1, -21, -146, - 133, -208 and -499 are specifically associated with acute myocardial infarction and acute coronary syndrome in humans.^4, 8^ MicroRNAs -499, -1-3 and -21 may identify patients with acute coronary syndrome even more robustly than high-sensitivity troponin.^9^ Thus, profiling circulating microRNA in patients undergoing elective non-cardiac surgery may provide genomic signatures that add mechanistic insight into perioperative myocardial injury. ^10, 11^

In this study, we hypothesised that if perioperative myocardial injury and acute coronary syndrome share pathophysiological mechanisms, then microRNA profiles should be similar in both. We therefore quantified levels of expression of microRNA in samples in serum and plasma extracellular vesicles obtained before and after non-cardiac surgery in patients who developed perioperative myocardial injury, compared to matched controls.

## Methods

### Study Populations

This was a nested case control study of samples obtained from UK patients enrolled in two separate studies of myocardial injury in non-cardiac surgery. Both studies were conducted in accordance with the principles of the Declaration of Helsinki and the Research Governance framework. The Measurement of exercise tolerance before surgery (METS) study was approved by the UK Medical research ethical committee (MREC: 13/LO/0135). 620 patients were recruited into the UK arm of the (METS) study cohort between 1^st^ March 2013 and 25^th^ March 2016.^12^ The second study, approved by the UK Medical research ethical committee MREC:16/LO/06/35, recruited 189 patients at University College London Hospital from 5^th^ October 2016 to 14^th^ January 2019.^13^ The protocol, including original inclusion/exclusion criteria have been published but are also provided in Supplementary data.

### Study 1: METS serum microRNA study

#### Inclusion/exclusion criteria

Inclusion criteria: Perioperative myocardial injury was defined as troponin I (TnI) >99^th^ centile (>0.04 ng.ml^-1^; Troponin-I Centaur CP assay, Siemens, UK). Patients with TnI>99^th^ centile after surgery were matched with patients in whom TnI remained ≤99^th^ centile after surgery. Samples were matched by age, sex, Revised Cardiac Risk Index Score and by day after surgery. All patients with confirmed PMI had an electrocardiogram reviewed by an independent adjudication committee We excluded patients with clinically confirmed myocardial infarction and/or pulmonary embolus, those receiving renal replacement therapy and participants with elevated troponin-I (TnI) >99^th^ centile before surgery (Figure 1).

**Figure 1:**
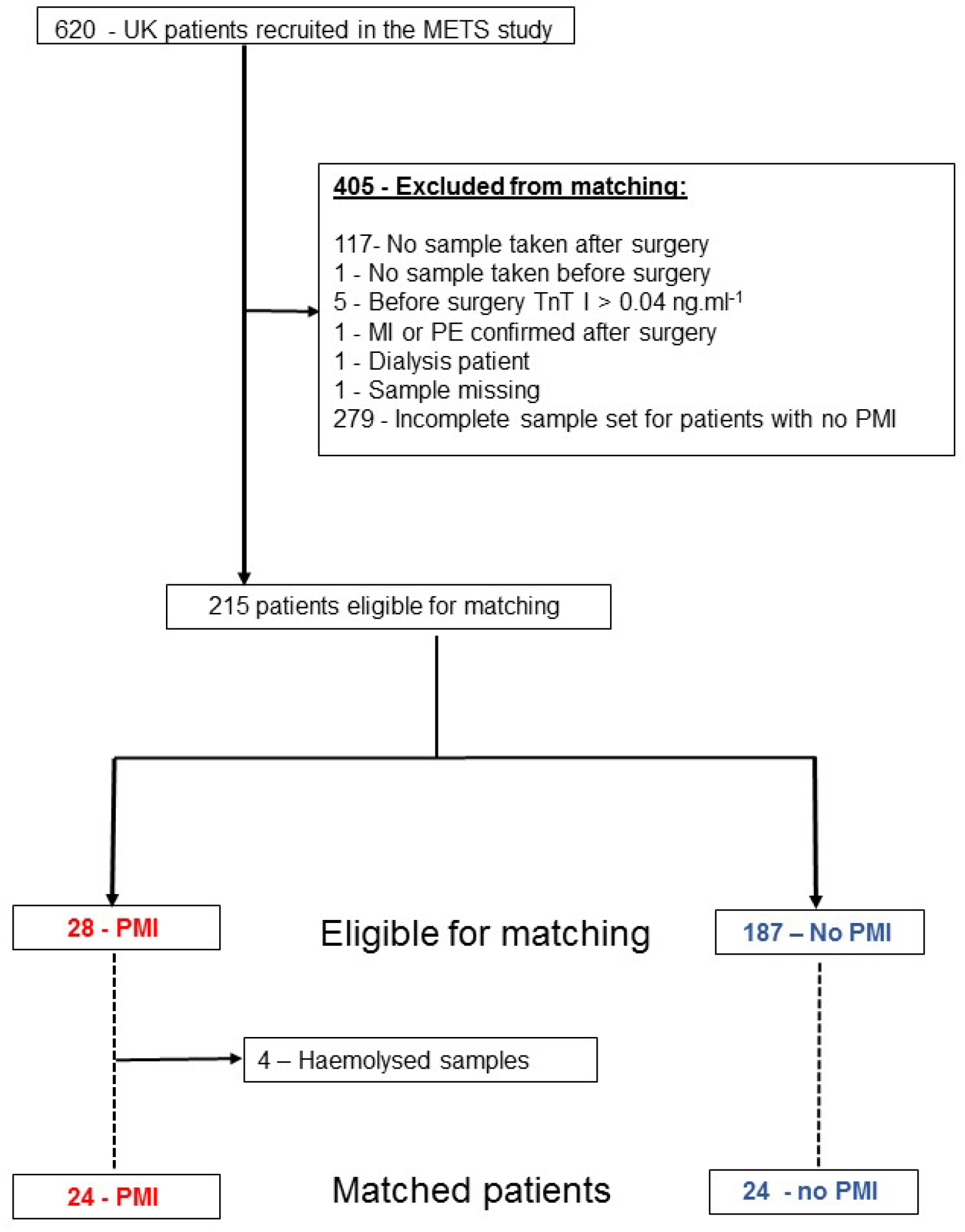
METS study 1 flow diagram, illustrating patient/sample selection.

#### METS study sample preparation

Samples were obtained once before surgery and then again on each day after surgery, until postoperative day 3. 10ml whole blood was collected in a plain tube, allowed to clot at room temperature before centrifugation (3000 rpm for 10 minutes). Serum was then aliquoted into two 2ml RNA-free tubes. The two aliquoted samples were then frozen at −70°C and subsequently used separately for troponin quantification and microRNA analysis.

#### METS study microRNA Isolation

Detailed methods and references are provided in Supplementary data. In brief, 200μl serum samples were screened for any obvious red discolouration (haemolysis) before isolation using the miReasy Serum/Plasma Advanced Kit (Qiagen, Denmark) in accord with manufacturer instructions. For quality control, 1μL of spike-in mixture containing UniSp2, UniSp4 and UniSp5 synthetic microRNAs (RNA Spike-in kit, Qiagen) was added to each sample (Supplementary data). 20uL RNA was isolated using RNeasy UCP MiniElute columns and stored at −20°C for subsequent batched analysis.

#### Circulating microRNAs

We selected microRNA-1, 21, 146, 133, 208 and 499 as established reported microRNA signatures for ACS (Supplementary table). Meta-analysis of microRNA discovery data has highlighted these microRNAs as potential biomarkers of myocardial infarction^14^. The isoforms selected were chosen from the clinical studies that had isolated microRNA from plasma/serum. The nomenclature was crosschecked using the miRBase database^15^ (Supplementary Material).

#### Quantitive RT-PCR

Detailed methods are provided in Supplementary Material. 2uL of RNA was reversed transcribed in a 10uL final reaction volume using the miRcury LNA RT kit. In accord with manufacturer recommendation, serum microRNA amounts are standardised by starting sample volume rather than RNA concentration. The reactions were performed in a Primus 96 plus thermal cycler (MWG Biotech, Ebersberg, Germany) for 60 minutes at 42°C, followed by incubation (inactivation of reaction) at 95°C for 5 min and then allowed to cool at 4°C. cDNA was then stored at −20°C. For qualitative PCR cDNA was diluted (1:40) and added to mastermix before plating on miRNA PCR custom made plates containing pre-designed miRCURY LNA miRNA PCR Assays in triplicate (Supplementary Data). Amplification was performed using the Applied Biosystem StepOneplus RT-PCR system (Applied Biosystems, California, USA). Cycle quantification (Cq) values were determined using the second derivative method (Applied Biosystem StepOneplus software, California, USA).

#### Quality control

Cq values were calibrated using an interplate calibrator control assay, UniSp3 pre-aliquoted on to each plate. The calibrated Cq data generated from the spike controls was used to compare each sample and identify outlier samples to be excluded from analysis. If the Cq values of the technical control produce similar values for each sample, isolation efficiency across all samples can be considered consistent. For this study, UniSp5 was used as the isolation technical control. If the Cq value was ≥37, then the sample and patient was flagged as an outlier and excluded from analysis. The synthetic RNA, UniSp6 was added to the RT reaction prior to cDNA synthesis. The outlier Cq values for UniSp6 were used to evaluate RT efficiency. Both hsa-miR-23a and hsa-miR-451were used to detect haemolysis, which interferes with miR detection ^16^. Samples that breached published limits of detection for haemolysis were excluded. Amplification efficiency was calculated (LinRegPCR software, Version 2017.1; Heart Failure Research Centre, Amsterdam, Netherlands)^17^ and individual melt curves for each PCR reaction were inspected (Supplementary data).

#### Cq normalisation and microRNA expression

The Qiagen miRcury LNA miRNA Serum/Plasma Focus Panel was used to determine two reference gene(s) for this study as recommended by consensus guidelines, since there are no universal reference microRNA defined for microRNA expression studies. ^18^ The focus panel has 179 microRNA assays commonly found in the serum and plasma. The NormFinder algorithm ^19^ was used to determine the most suitable reference microRNA (Supplementary material). MicroRNA miR-152-3p and hsa-miR-361-5p had the lowest combined stability value (0.077) and selected as reference microRNA. Expression levels were compared using the relative Cq method, where microRNA Cq values were normalised to the geometric mean of the reference microRNA Cq values to give ΔCq values. The ΔCq values were converted to a fold change (FC) by applying the formula of 2^−ΔCq. 20^

#### Pathway analysis

We used DNA Intelligent Analysis (DIANA)-miRPath v.3.0 pathway analysis software^21^ to assess in an unbiased manner which transcriptomic processes may be altered by microRNAs (microT-CDS v5.0).

### Study 2: microRNA expression levels in extracellular vesicles

#### Extracellular vesicles samples

To examine microRNA content in extracellular vesicles, a separate cohort of patients was recruited to prepare larger volumes of plasma which required specific different preparation to the samples obtained in METS. Plasma samples were prepared from 11 patients who underwent elective non cardiac surgery.^13^ Perioperative myocardial injury was defined as plasma high sensitivity Troponin T assay ≥99^th^ centile (≥15 ng.L^-1^) within 2 days of surgery (Cobas assay. Roche Diagnostics, Mannheim, Germany). 48h after surgery whole blood was collected into EDTA anticoagulant bottles, centrifuged at 3000 RPM for 10 minutes (Cepr-AL6 Flexicool Rota, Capricorn Laboratory Equipment, UK). The plasma layer was sterile filtered (0.8μm membrane filter, Starlabs, Hamburg, Germany) and transferred to Eppendorf tubes. The samples were immediately stored at −80°C until batch processing. At the same time blood samples were collected, an ECG was reviewed independently to ensure no patient with myocardial infarction was included.

#### microRNA isolation from extracellular vesicles

Plasma samples were shipped to Exiqon (Vedbaek, Denmark) to isolate microRNA in extracellular vesicles and exosomes for next generation sequencing, masked to troponin values and clinical details. Quality control checks ensured high isolation efficiency and lack of haemolysis.^16^ (Supplementary Material) Library preparation was done using the NEBNext® Small RNA Library preparation kit (New England Biolabs, Massachusetts, USA). The library pool was sequenced on a NextSeq500 sequencing instrument (Illumina, San Diego, USA). Raw data was de-multiplexed and FASTQ files for each sample was generated using the bcl2fastq software (IIllumina, San Diego, USA). The MicroRNA sequence reads (FASTQ) files were then mapped to Homo sapien genome (GRCh37) and the microRNA sequence database, MiRBase V20, to generate MicroRNA sequence counts using Bowtie2 (Version 2.2.4.) software.^22^ MicroRNA counts were normalised for library size using Trimmed mean of M (TMM) method and differential expression of TMM analysis performed using EdgeR statistical software package.^23^ Differentially expressed microRNA were used to perform Gene ontology enrichment analysis using topGO.^24^

#### Statistical Analysis

Clinical characteristics of the patients were stratified according to presence, or absence, of troponin elevation >99^th^ centile values. Categorical data are summarised as absolute values (percentage). Continuous data are presented as mean (SD), or median (IQR), unless stated otherwise. ΔCq values were checked for normality using the Shapiro–Wilk test. For microRNA assays that were expressed in >95% across all samples, ΔCq values were analysed using a multi-level (mixed error-component) model. The levels examined were; presence/absence of perioperative myocardial injury (>99^th^ centile) and sampling time (before, or after, surgery).

Individual comparisons between groups were calculated using post-hoc Bonferroni tests. For microRNA expressed in <95% samples, McNemar Chi-square analysis was performed. P<0.05 was considered statistically significant. All statistical analyses were undertaken using NCSS 12 (Kaysville, UT, USA). Data have been deposited in NCBI’s Gene Expression Omnibus^25^ and are accessible through GEO Series accession number GSE144777.

#### Sample Size Calculation

Receiver operating character curve analysis has demonstrated that miR-499, miR-1-3 and miR-21 can be used to identify patients with acute coronary syndrome, area under the curve (AUC) = 0.89. ^9^ If these microRNA were present and/or elevated in patients at the time of detecting perioperative myocardial injury, >11 patients would be required to detect a difference in relative expression serum microRNA after surgery (α=0.01; 1-β=0.9). The sample size calculation was performed using the pROC package (R software, v3.4.3).^26^

## Results

### Cohort 1: METS serum microRNA study

#### Patient characteristics

Patients recruited from METS who developed myocardial injury (median TnI: 0.08ng.ml^-1^ (interquartile range: 0.05-0.13ng.ml^-1^)) were clinically similar to patients in whom troponin remained within normal limits (Table 1).

**Table 1:** Patient characteristics for METS study cohort. Data is presented as mean with standard deviations (SD) for parametric data and as median (25th-75th interquartile range) for non-parametric data. Frequencies are presented with percentages (%). Age is rounded to the nearest year. Perioperative myocardial injury (PMI) defined was defined as troponin I (TnI) >99^th^ centile (>0.04 ng.ml^-1^) after surgery. eGFR: estimated glomerular filtration rate. RCRI: Revised cardiac risk index score. ACE-I: Angiotensin converting enzyme inhibitor. ARB: angiotensin receptor blocker. Other units as indicated. ^#^Previous diagnosis of asthma, reactive airways disease, chronic obstructive lung disease, chronic bronchitis, or emphysema. *Indication for surgery was for treatment of cancer. Statistical analysis using paired-sample t-test or Wilcoxon-Signed rank test for continuous data and Chi-Squared test for categorical data.

|  | Whole cohort<br>n=48 | No PMI<br>n=24 | PMI<br>n=24 | *p-value* |
| --- | --- | --- | --- | --- |
| Troponin level ng ml<sup>-1</sup> |  | ≤0.04 | 0.08 (0.05-0.13) | <0.001 |
| **Clinical Characteristics** |  |  |  |  |
| Age (yr) | 67 (61-75) | 67 (60-74) | 66 (61-76) | 0.154 |
| Male sex (n;%) | 22 (45.8%) | 11 (45.8%) | 11 (45.8%) | 1.000 |
| Body Mass Index (kg m<sup>-2</sup>) | 27 (5) | 27(5) | 27 (5) | 0.900 |
| Before surgery eGFR (ml.min<sup>-1</sup>.1.73m<sup>2</sup>) | 87 (76-96) | 84 (72-94) | 90 (79-103) | 0.192 |
| Haemoglobin (g dl<sup>-1</sup>) | 130 (22) | 128 (16) | 131 (26) | 0.520 |
| **Co-morbidities (n; %)** |  |  |  |  |
| RCRI ≥2 | 4 (8.3%) | 2 (8.3%) | 2 (8.3%) | 1.000 |
| Coronary artery disease | 5 (10.4%) | 3 (12.5%) | 2 (8.3%) | 0.637 |
| Congestive heart failure | 0 (0.0%) | 0 (0.0%) | 0 (0.0%) | - |
| Cerebrovascular disease | 1 (2.1%) | 1 (4.2%) | 0 (0.0%) | 0.312 |
| Diabetes Mellitus | 10 (20.8%) | 5(20.8%) | 5 (20.8%) | 1.000 |
| Peripheral vascular disease | 1 (2.1%) | 0 (0.0%) | 1 (4.2%) | 0.312 |
| Hypertension | 24 (50.0%) | 11(45.8%) | 13 (54.2%) | 0.564 |
| Obstructive lung disease<sup>#</sup> | 5 (10.4%) | 3 (12.5%) | 2 (8.3%) | 0.636 |
| Significant malignancy* | 34 (70.8%) | 18 (75.0%) | 16 (66.7%) | 0.524 |
| **Preoperative medications** |  |  |  |  |
| Beta blocker | 6 (12.5%) | 4 (16.7%) | 2 (8.3%) | 0.383 |
| Calcium channel blocker | 5 (10.4%) | 3 (12.5) | 2 (8.3%) | 0.637 |
| ARB/ACE-I | 19 (39.5%) | 8 (33.3%) | 11 (45.8%) | 0.376 |
| Aspirin | 8 (16.7%) | 4 (16.7%) | 4 (16.7%) | 1.000 |
| **Surgical Procedure type (n; %)** |  |  |  |  |
| Intraperitoneal or retroperitoneal | 30 (62.5%) | 17 (70.8%) | 13 (54.2%) | 0.223 |
| Urology or gynaecological | 15 (31.3%) | 6 (25.0%) | 9 (37.5%) | 0.350 |
| Orthopaedic | 3 (6.2%) | 1 (4.2%) | 2 (8.3%) | 0.551 |
| **Anaesthesia Type (n; %)** |  |  |  |  |
| General Anaesthesia alone | 24 (50.0%) | 13(54.2%) | 11 (45.8%) | 0.564 |
| General + regional anaesthesia | 24 (50.0%) | 11 (45.8%) | 13 (54.2%) | 0.564 |

#### Plasma circulating microRNA expression

Circulating microRNA expression levels increased for hsa-miR-1-3p (mean fold-change:3.99 (95%CI: 1.95-8.19); F_[1,47.5]_ =24.16; p<0.001) and hsa-miR-133-3p (fold-change:5.67 (95%CI: 2.94-10.91); F_[1,42]_ = 31.13; p<0.001) after surgery, but were independent of elevated troponin levels after surgery (mixed model level accounting for presence/absence of PMI: p>0.05). Expression levels of hsa-miR146a-5p decreased after surgery only in patients with elevated troponin after surgery (F_[1,34.5_]=6.43, p=0.03). There was no change in expression for hsa-miR-21-5p after surgery (Figure 2). In patients with troponin elevation after surgery, 91% (20/22) of the samples detected hsa-miR-208b-3p, compared with 50% (12/24) with troponin levels below the 99^th^ centile after surgery (odd ratio (OR):10.00 (95%CI:1.90-52.55); p<0.01)). hsa-miR-499a-5p was also detected in a higher proportion of samples from patients with elevated troponin levels after surgery (19/22 vs 13/24; OR:5.36 (95% CI 1.32-24.17); p<0.05) surgery. (Supplementary Material; Figure 1)

**Figure 2:**
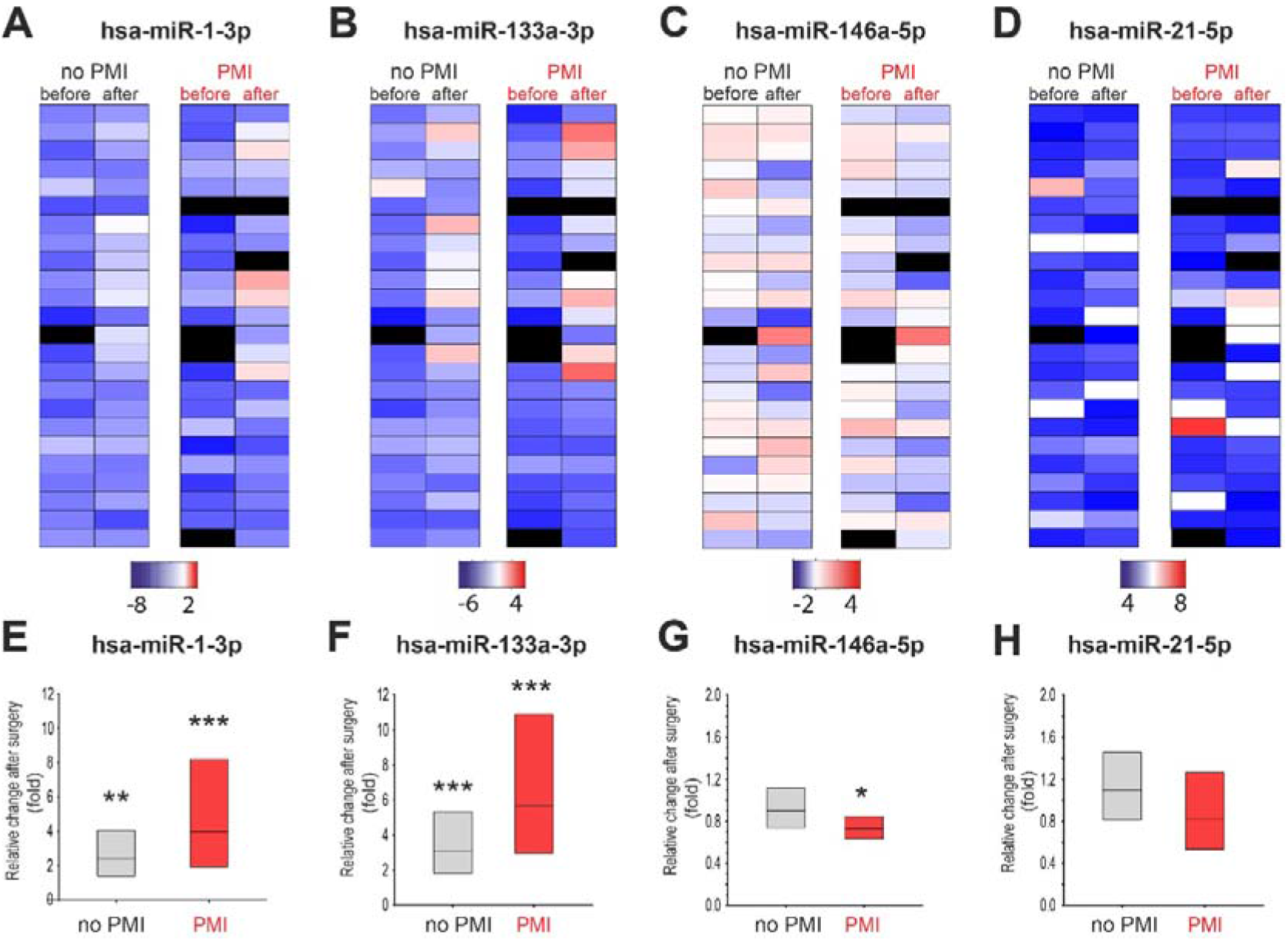
Heatmaps showing relative expression levels (Fold change) of detectable serum microRNA before and after non-cardiac surgery, in relation to the development of myocardial injury. **A-D:** Heatmap showing relative serum expression levels for each matched sample using the relative Cq method. MicroRNA Cq values were normalised to the geometric mean of the reference microRNA Cq values to give ΔCq values. The ΔCq values were converted to a fold change (FC) by applying the formula of 2^−ΔCq^. Bars below each microRNA heatmap indicate range of fold-change expression (log_2_ scaled). Black bars denote sample that failed strict quality control threshold. **E-H:** Relative change in MicroRNA expression levels after surgery. Bars are mean fold change with 95% confidence intervals. P-values refer to comparison; before surgery vs after surgery (analysed by mixed-model repeated measures and Bonferroni test); * p<0.05, ** p <0.01 **p<0.001. PMI: perioperative myocardial injury (defined as troponin I (TnI) >99th centile (>0.04 ng.ml^1^).

#### Pathway analysis

Nine signalling pathways were identified by DIANA in-silica pathway analysis (false-discovery rate <0.01) for microRNA that changed after surgery (Figure 3A). This agnostic analysis approach highlighted the involvement of adrenergic signalling in cardiomyocytes (hsa04261; p=0.005) and arrythmogenic right ventricular cardiomyopathy (hsa05412; p= 0.006). All microRNAs that were elevated in serum after surgery (n=4) regulated expression of SLC8A1, a gene that controls calcium overload through the NCX1 antiporter membrane protein by removing calcium from cells (Figure 3B). Subunits of protein phosphatase (which regulates cardiac excitability through dephosphorylation of cardiac ion channels), angiotensin receptors and the ATP sarcoplasmic/endoplasmic reticulum calcium pump were also identified as targets of microRNAs that changed after surgery. (Figure 3B)

**Figure 3:**
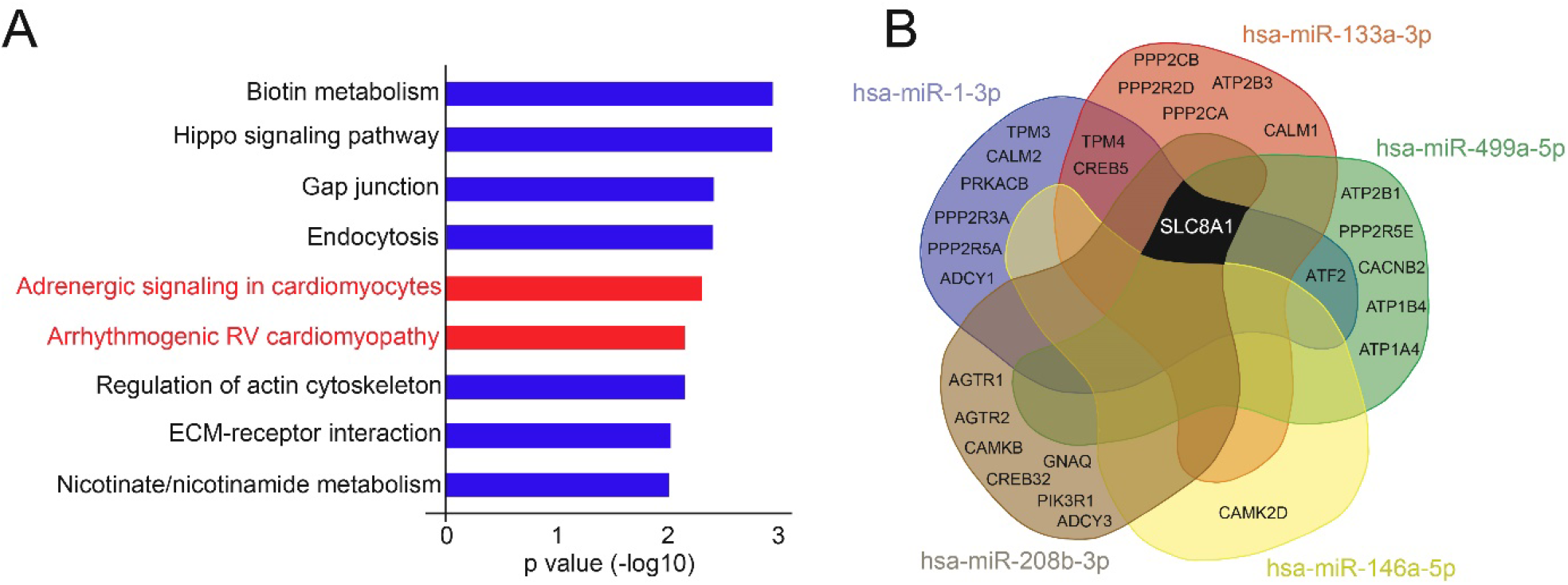
Bioinformatic analysis identifying pathways and genes regulated by serum microRNAs that change after non-cardiac surgery. A. Signalling pathways (false-discovery rate threshold p value <0.01), as derived from DIANA mir-Path v3.0, which bioinformatically links microRNA with their regulatory roles in biological and signalling pathways. Cardiac-specific pathways are highlighted in red. B. Shared genes regulated by serum microRNA identified as changing after non-cardiac surgery. The four microRNAs identified as changing after surgery regulate mRNA expression of SLC8A1 (NCX1 sodium-calcium exchanger) and subunits of the protein phosphatase enzyme.

### Cohort 2: microRNA expression levels in extracellular vesicles

#### Patient characteristics

Characteristics of the 11 patients from whom samples were obtained 48 hours after surgery are presented in Table 2. The median (IQR) troponin-T values were 19 (17-23) ng.L ^-1^ in patients with troponin elevation, compared with patients in whom plasma troponin remained lower than the 99th centile value (median: 6 (5-9) ng.L^-1^; p=0.006).

**Table 2:** Patient characteristics of X-MINS study cohort. Data is presented as mean with standard deviations (SD) for parametric data and as median (25th-75th interquartile range) for non-parametric data. Frequencies are presented with percentages (%). Age is rounded to the nearest year. Perioperative myocardial injury was defined as plasma high sensitivity Troponin T Assay ≥99^th^ centile (≥15 ng.L^-1^) within 2 days of surgery. eGFR: estimated glomerular filtration rate. RCRI: Revised cardiac risk index score. ACE-I: Angiotensin converting enzyme inhibitor. ARB: angiotensin receptor blocker. Other units as indicated. ^#^Previous diagnosis of asthma, reactive airways disease, chronic obstructive lung disease, chronic bronchitis, or emphysema. *Indication for surgery was for treatment of cancer.

|  | Whole cohort | No PMI<br>n=5 | PMI<br>n = 6 |
| --- | --- | --- | --- |
| Troponin level ng L<sup>-1</sup> |  | 6(5-9) | 19 (17-23) |
| **Clinical Characteristics** |  |  |  |
| Age (yr) | 70 (64-76) | 68(62-71) | 74 (68-81) |
| Male sex (n;%) | 3 (27.3%) | 1 (20%) | 2 (33.3%) |
| Body Mass Index (kg m<sup>-2</sup>) | 31 (8) | 30 (7) | 32 (9) |
| Before surgery eGFR (ml.min<sup>-1</sup>.1.73m<sup>2</sup>) | 83(65-90) | 77 (66-89) | 87(49-90) |
| Haemoglobin (g L<sup>-1</sup>) | 12.5 (2.1) | 12.7 (1.7) | 12.4 (2.5) |
| **Co-morbidities (n; %)** |  |  |  |
| RCRI $\geq 2$ | 2 (18.2%) | 1 (20%) | 1 (16.7%) |
| Coronary artery disease | 0 (0.0%) | 0 (0%) | 0 (0%) |
| Congestive heart failure | 0 (0.0%) | 0 (0%) | 0 (0%) |
| Cerebral vascular disease | 2 (18.2%) | 1 (20%) | 1 (16.7%) |
| Diabetes mellitus | 2 (18.2%) | 0 (0%) | 2 (33.3%) |
| Peripheral vascular disease | 1 (9.1%) | 1 (20%) | 0 (0%) |
| Hypertension | 6 (54.5%) | 3 (60%) | 3 (50%) |
| Obstructive lung disease<sup>#</sup> | 3 (27.3%) | 1 (20%) | 2 (33.3%) |
| Significant malignancy<sup>*</sup> | 7 (46.6%) | 2 (40%) | 5 (83.3%) |
| **Preoperative medications** |  |  |  |
| Beta blocker | 2 (18.2%) | 0 (%) | 2 (33.3%) |
| Calcium channel blocker | 1 (9.1%) | 0 (0%) | 1 (16.7%) |
| ARB/ACE-I | 4 (36.4%) | 2 (40%) | 2 (33.3%) |
| Aspirin | 2 (18.2%) | 2 (40%) | 0 (0%) |
| **Type of surgery (n; %)** |  |  |  |
| Intraperitoneal or retroperitoneal | 2 (18.2%) | 1 (20%) | 1 (16.7%) |
| Urology/gynaecological | 5 (45.4%) | 2 (40%) | 3 (50%) |
| Orthopaedic | 4 (36.4%) | 2 (40%) | 2 (33.3%) |
| **Type of anaesthesia (n; %)** |  |  |  |
| General Anaesthesia alone | 4 (36.4%) | 2 (40%) | 2 (33.3%) |
| General + regional anaesthesia | 7 (63.6%) | 3 (60%) | 4 (66.7%) |

#### Next generation sequencing

A mean of 9.8 million read were obtained for each sample. Mean mapping rate to the genome and microRNA annotation was 62.9%. Mapping to miRbase, with normalisation for differences in the sequences depth within each sample, revealed 345 unique microRNA within all samples where Tags per million (TPM) ≥1 and 164 unique microRNAs where TPM ≥10. Thirty-two microRNAs were differentially expressed in patients with troponin elevation after surgery (Figure 4A). After correcting for false discovery rate <0.05, three microRNAs remained differentially expressed (Figure 4B). The top two biological processes identified through Gene Ontology were associated with cardiac muscle physiology. (Figure 4C; Supplementary material).

**Figure 4:**
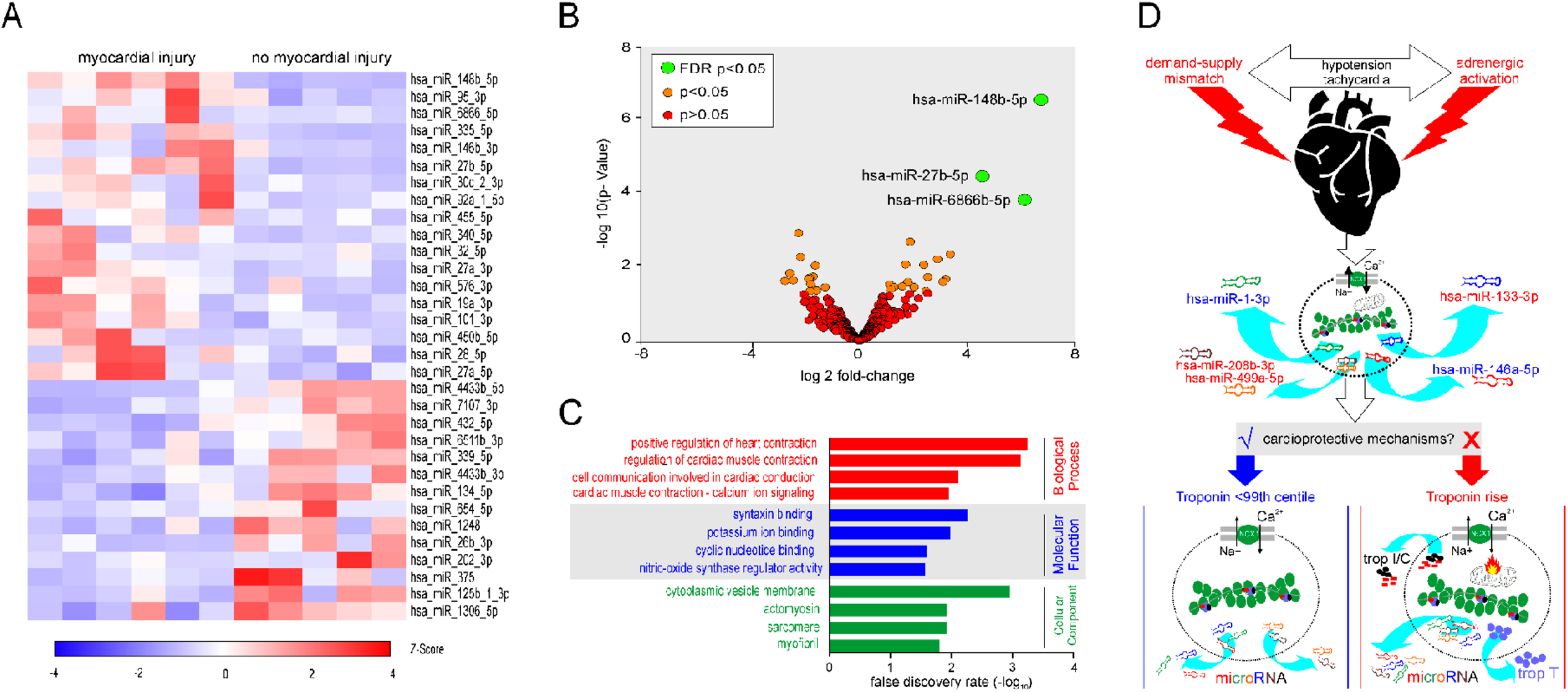
Next generation sequencing of microRNAs detected in extracellular vesicles. **A**. Heat map showing all differentially expressed microRNA (P<0.05), comparing patients with PMI (hsTnT >14ngL^-1^) versus no troponin elevation. Values are Trimmed mean of M values. (Z score transformation; red indicates higher expression levels). **B**. Volcano plot of microRNAs identified in all samples. Green dots denote differentially expressed microRNA with adjusted p-value (FDR; false discovery rate) <0.05. Orange dots denote differentially expressed microRNA with p-value <0.05 and red dots denote microRNA that were not differentially expressed. **C**. GO terms that were significantly enriched among the differentially expressed microRNA, comparing patients with PMI (hsTnT >14ngL^-1^) versus no troponin elevation. **D**. Refined hypothesis of mechanisms contributing to perioperative myocardial injury, summarising chief microRNA findings and their potential roles in cardioprotection. Blue coloured microRNA species have been associated with cardioprotection, while red coloured microRNAs promote cardiac injury in the setting of ischaemia/reperfusion injury. Trop I-troponin I; Trop C-troponin C; Trop T-troponin T. Adrenergic stress is a common feature of the perioperative period, leading to the release of microRNA from cardiac cells, which may be cardiomyocytes, fibroblasts or other cell types. In individuals at highest risk of elevated troponin after surgery (e.g. elderly, patients with cardiac failure, diabetes mellitus, renal disease), pre-exisiting (preoperative), or acquired, loss of cardioprotective signalling mechanisms promotes the likelihood of calcium overload from adrenergic stress, resulting in mitochondrial dysfunction, bioenergetic compromise and myocardial injury manifest clinically by the measurement (at low levels) of troponin T in plasma/serum.

## Discussion

Our data provide further mechanistic insight into perioperative myocardial injury by quantifying circulating microRNAs from samples obtained serially in patients undergoing non-cardiac surgery. This exploratory study demonstrates that microRNAs are readily detected in serum and extracellular vesicles obtained from patients undergoing non-cardiac surgery. Using technical spike-in quality controls and standardised, reproducible sampling protocols, we report that circulating levels of cardiac-specific microRNAs that increase after ACS^9, 27^ were also elevated after non cardiac surgery. MicroRNA expression levels were independent of troponin levels, suggesting strongly that several microRNA signatures described in the acute coronary syndrome literature reflect a generalised stress/injury response. In support of our circulating microRNA data, agnostic identification of microRNA expression in extracellular vesicles (which, again, was also masked to serial troponin values) revealed pathways from differentially expressed microRNAs that predominantly regulate cardiac muscle contraction. Our findings are consistent with the activation of G protein-coupled receptor signaling mediators, which have an established role through β-arrestins in regulating processing of microRNAs.^28^ GPCR-mediated stress signalling appears likely to be a dominant factor that contributes to troponin elevation after surgery, since microRNA findings described for myocardial ischaemia/infarction were not recapitulated by our agnostic bioinformatic analyses. Taken together, microRNA data from both plasma extracellular vesicles and serum suggest that a cardiac-stressor phenotype is common amongst patients after non-cardiac surgery, regardless of the development of troponin release and myocardial injury.

Several microRNA have been reported as specific biomarkers for acute coronary syndrome, including miR-499, miR-1, miR-133a/b, and miR-208a/b. In a prospective single-centre study, levels of cardiac miRNAs (miR-1, -208a and -499), miR-21 and miR-146a increased in 106 patients diagnosed with ACS, even in patients with initially negative high-sensitive troponin or symptom onset <3 h.^9^ The combination of miR-1, miR-499 and miR-21 increased the diagnostic value in all suspected ACS patients, over and above that for hs-troponin T. ^9^ However, in contrast to microRNA studies in acute coronary syndrome, we found that troponin elevation after surgery was not related to ACS-specific microRNAs did including key players such asmiR-1 and miR-133a. In the case of hsa-miR-146a-5p, expression levels were decreased in patients who developed perioperative myocardial injury, in direct contrast to elevated circulating levels reported after myocardial infarction, ^9, 29^ MicroRNA-208, a cardiac specific microRNA that regulates cardiac myosin heavy chain expression, ^30^ increases in patients after acute myocardial infarction ^31^ but we detected miR-208a in only ∼10% patients. Similarly, the low level of detection rates for miR-208b in our study are in contrast to 208b expression levels correlating with troponin levels ^32^ including after acute myocardial infarction ^29^. Although hsa-miR-499a-5p was detected in a higher proportion of patients who developed perioperative myocardial injury, the detection rates for both miR-208b-3p and miR499a-5p we found are consistent with low detection rates reported in the literature^9, 27^ and low constitutive levels in plasma/serum^33^.

Our two independent studies analysing microRNA derived from separate sources (serum, extracellular vesicles) suggested common mechanistic targets. Agnostic bioinformatic analyses highlighted microRNA changes in both serum and extracellular vesicles that converge on established gene transcription targets that regulate cardiomyocyte cell calcium homeostasis,^34^ predominantly involving the sodium-calcium exchanger (NCX1) which is encoded by the SLC8A1 gene. ^35^ Calcium, which enters cardiac myocytes through voltage-dependent Ca(2+) channels during excitation, is extruded from myocytes primarily by NCX1.^28^ Ischemia/reperfusion reverses the sodium/calcium exchange mechanism, leading to an increase in intracellular calcium concentration, mitochondrial dysfunction and cardiomyocyte death. However, sustained stress also inhibits NCX1 activity, an effect that requires corticosteroid release in concert with adrenergic activation.^36^

Reversible protein phosphorylation is essential for excitation-contraction coupling, Ca2+ handling, cell metabolism, myofilament regulation, and cell-cell communication.^37^ ∼90% of dephosphorylation events are catalysed by just three phosphatases: PP1, PP2A and PP2B (calcineurin), all of which are regulated by microRNA we identified as changing after surgery. We identified multiple potential PP2A regulatory subunits potentially related to microRNA changes after surgery, which likely reflects the diversity of expression between cardiac cell types, subcellular domains, and disease phenotypes.^37^ Cardiac myosin binding protein-C (cMyBP-C) and the inhibitory subunit of troponin TnI are also dephosphorylated by both PP1 and PP2A. Targeting of PP2A to TnI is dependent upon the PP2A regulatory subunit B56α, which is lost following β-adrenergic stimulation. Aberrant regulation of PP2A signaling is a dominant feature in experimental sympathetic-mediated tachycardia and generation of reactive oxygen species.

In the universal presence of a stressor phenotype after surgery, elevations in plasma troponin suggest that loss of cardioprotective mechanisms promote myocardial injury (Figure 4D). An aberrant inflammatory response, as indicated by the decline in expression levels of the anti-inflammatory microRNA hsa-miR-146a-5p, is a plausible mechanism. By inhibiting expression of IRAK1, TRAF6 and NFκβ, hsa-miR-146a-5p suppresses the production of pro-inflammatory cytokines^38^ which promote myocardial injury.^39^ Since inflammation is strongly associated with the risk of developing perioperative myocardial injury,^3^ reduced expression of hsa-miR-146a-5p indicates that dysregulation of inflammation after surgery may contribute to myocardial injury. Furthermore, loss of cardioprotective vagal tone,^40^ which occurs rapidly in many patients during non-cardiac surgery,^13^ is associated with troponin elevation after non-cardiac surgery. Experimental preservation of vagal activity after myocardial ischaemia maintains normal calcium handling by restoring the protein and mRNA levels of sarcoplasmic reticulum Ca^2+^ ATPase (SERCA2a), NCX1 and phospholamban.^41^ The main strength of our study was investigating two separate populations in which we could obtain samples using optimised protocols that enabled microRNA detection in serum and plasma extracellular vesicles to address our overarching hypothesis. Moreover, we compared serial changes in microRNA masked to troponin levels before and after surgery. Reference microRNA were used for normalisation of Cq values. For microRNA that were detected in all samples, the Cq values were consistent with values reported in the literature.^29^ In fact, the detection threshold in clinical studies using the same microRNA isolated from serum have used much higher threshold i.e Cq <50.^27^ The utilisation of the spike-in microRNA at isolation, reverse transcription and PCR stage of the experiments allowed for identification of outliers and minimised technical variation within the experiments. A further strength of this work is that the amplification efficiencies are reported in keeping with MIQE guidelines. The low efficiency values for hsa-miR-208b-3p and hsa-miR499a-5p are consistent with reported low detection rates for both.^42^ Our limited targeting of circulating serum microRNAs that are established genomic biomarkers for acute coronary syndrome and myocardial infarction clearly cannot exclude other novel species that may be involved in perioperative myocardial injury. However, the purpose of this initial exploratory study was to investigate whether established ischaemic and/or thrombotic microRNA signatures are present after troponin elevation in non-cardiac surgery.

In summary, expression levels of circulating microRNA that are elevated in acute coronary syndromes also increase after non-cardiac surgery, but this pattern does not appear to be specific for patients with perioperative myocardial injury (as defined by high-sensitivity troponin). MicroRNA changes after non-cardiac surgery appear to occur as a feature of generalised cardiac stress, suggesting that myocardial injury is promoted by deficient cardioprotective mechanisms that prevent troponin release from otherwise injured cardiomyocytes.

## Data Availability

Data have been deposited in NCBI's Gene Expression Omnibus and are accessible through GEO Series accession number GSE144777.

## Author Contributions

Study design: GLA, SMM.

Study conduct: SMM, TEFA, AGDA, AR, GM, RCMS, DB, BHC, DNW, RMP, GLA

Experiments: SMM, AGDA, GLA

Data analysis: SMM, GLA

Drafting/revising manuscript: GLA, SMM.

Critical review: all authors.

## Declarations

GLA is an Editor for Intensive Care Medicine Experimental and British Journal of Anaesthesia. GLA has undertaken consultancy work for GlaxoSmithKline; TEFA is a member of the associate editorial board of British Journal of Anaesthesia; there are no other relationships or activities that could appear to have influenced the submitted work.

